# Norwegian general population normative data for the EORTC questionnaires; the core QLQ-C30, the sexual health questionnaire QLQ-SHQ22, and sexual domains of the QLQ-BR23/BR45

**DOI:** 10.1101/2023.02.25.23286292

**Authors:** R.E Åsberg, M Nilsen, M.J Hjermstad, K.V Reinertsen, J Karlsen, G.F Giskeødegård, R.J. Reidunsdatter

**Affiliations:** Department of Circulation and Medical Imaging, Faculty of Medicine, and Health Sciences, NTNU; Department of Social Work, Faculty of Social and Educational Sciences, NTNU; K.G. Jebsen Center for Genetic Epidemiology, Department of Public Health and Nursing, Faculty of Medicine and Health Sciences, NTNU; Department of Surgery, St. Olavs University Hospital; Regional Advisory Unit for Palliative Care, Dept. of Oncology, Oslo University Hospital / European Palliative Care, Research Centre (PRC) and Institute of Clinical Medicine, University of Oslo, Oslo; Department of Oncology, Oslo University Hospital; National Advisory Unit for Late effects After Cancer Treatment, Department of Oncology, Oslo University Hospital, Oslo; Department of Oncology, St.Olav’s Hospital, Trondheim University Hospital, Trondheim, Norway; Department of Clinical and Molecular Medicine, Faculty of Medicine and Health Sciences, NTNU, Trondheim, Norway

**Keywords:** Health-Related Quality of Life, HRQOL, normative values, Norwegian general population, patient-reported outcomes, PROMs, EORTC QLQ-C30, QLQ-BR23, QLQ-BR45, QLQ-SH22, co-morbidity, sexual scales, cancer

## Abstract

**Objective:** The aim of this study was to provide gender-, age-, and morbidity-specific Norwegian general population normative values for the European Organisation for Research and Treatment of Cancer Quality of Life Questionnaires QLQ-C30, the sexual health questionnaire QLQ-SHQ22, and the sexual domains of the breast modules QLQ-BR23 and QLQ-BR45.

**Methods:** A random nation-wide sample stratified by gender and age groups (18–29, 30-39, 40–49, 50–59, 60–69 and ≥70 years) was drawn from the Norwegian National Population Register. Participants were notified through National online health services (HelseNorge) and by postal mail. The survey included sociodemographic background information, HRQoL assessed by the EORTC questionnaires, and morbidity by The Self-Administered Comorbidity Questionnaire. Multivariable linear regression was carried out to estimate the associations of age, sex, and morbidity with the EORTC scale and item scores.

**Results:** Of the 15,627 eligible individuals, 5.135 (33%) responded. Women and persons with morbidities reported lower functioning and higher symptom burden than men and persons without morbidities, respectively, on nearly all EORTC scales. Sex differences were most prominent for *Emotional Functioning, Pain, Fatigue*, and *Insomnia* (QLQ-C30), *Body Image, Sexual Functioning* (QLQ-BR23/45), *Importance of Sexual Activity, Libido*, and *Fatigue* (QLQ-SHQ22). The score differences between persons with and without morbidity were highly significant and largest among the youngest and middle-aged groups.

**Conclusion:** The present study is the first to provide normative values for the EORTC sexual health questionnaire QLQ-SHQ22 and the sexual subscales of the QLQ-BR23 and QLQ-BR45, for all separately in age groups by sex and morbidity.

## Introduction

Patient-reported outcome measures (PROMs) reflect the patients’ own perceptions of health-related quality of life (HRQoL) [1, 2]. The purpose of PROMs in oncology is to provide valuable information on how cancer and treatment affect short- and long-term HRQoL and thereby guide clinicians in patient-centered care [3-5]. PROMs are recognized as independent endpoints in clinical studies and health care research worldwide [6].

One of the most frequently used PROMs in oncology is the European Organization for Research and Treatment of Cancer Quality of Life Questionnaire-Core 30 (EORTC QLQ-C30) [5, 7]. This cancer specific core questionnaire is often supplemented by modules for specific diagnoses or conditions, e.g. the breast cancer modules QLQ-BR23 / QLQ-BR45 [8, 9].

Even though these questionnaires are validated and frequently used, meaningful and consistent interpretations of scores remain challenging in both research and clinical practice[10-12]. One approach is to compare changes in PROMs at the group or patient level using clinically significant differences [13]. For interpretation of EORTC scores obtained at certain time points, thresholds for high symptom scores or low functional scores have been utilized [14, 15], and for the core EORTC QLQ-C30, thresholds clinical importance of domains have recently been developed [16]. General population data provide estimates of HRQOL scores of the same age and sex as the patients, and thereby supporting the interpretation of PROMs in clinical practice and cancer studies [17-20].

Sexual health is an important aspect of HRQoL [21, 22], and sexual problems have a high prevalence of in cancer survivors [23]. Despite this fact, the only normative EORTC data on sexual health are from a Dutch normative study incorporating five sexual single items from the EORTC’s item bank [24]. So far sexual concerns have been covered by a few items in some the EORTC modules, e.g., the two breast cancer modules QLQ-BR23 and QLQ-BR45 [8, 9] Thus, EORTC has recently developed a standalone Sexual Health Questionnaire (QLQ-SHQ22) for a more comprehensive assessment of sexual health [25]. To date, no normative data on sexual health have been published from Scandinavia.

Perceived HRQoL varies by age and sex and during the life course [17, 20, 26-29], and further, poor health condition has a great negative impact on HRQOL [5, 19, 27, 30]. Hence, a valid assessment of morbidity should be included in the collection of normative data, and comorbidity should be accounted for in comparisons with cancer populations. As such, normative data may provide information about health issues that are probably due to the cancer or treatment or simply an effect of normal ageing, morbidities or sex differences [5, 19, 27, 31] Country-specific normative values have been conducted for European countries, included Norway in 1998 and 2007 [30, 32], showing national differences [5, 18-20, 24, 27-29, 31, 33-38]. Updated data are essential to display the current HRQoL in the Norwegian general population.

The primary aim of this study was to provide sex and age specific normative values from the Norwegian general population for HRQoL, including sexual health addressed by the EORTC questionnaires QLQ-C30, QLQ-SHQ22, and the sexual domains of QLQ-BR23 and QLQ- BR45.

## Methods

### Study procedure and participants

The study was designed as a nationwide electronic and postal cross-sectional survey. The web solution eFORSK (https://www.klinforsk.no/info/Informasjon), developed by the Central Norway Regional Health Authority IT department (HEMIT) and run by the Norwegian Health Network (NHN), was utilized for data collection. A pilot study including 15 participants was performed to test the comprehensibility of the survey and the usability of the digital platform (eFORSK) for data collection, and adaptations were carried out accordingly.

The Norwegian Tax Administration gave permission to draw a randomly selected sample (N=15.627) from the Norwegian National Population Register stratified by gender and age. This sample size was estimated to ensure sufficient sample sizes for age subgroups (18-29, 30–39, 40–49, 50–59, 60–69 and ≥ 70 years). The data extraction was executed by the national IT-company Evry (Evry.com). To increase the response rate, the randomly selected participants received a digital postcard informing them about the upcoming study a week ahead of the survey release. The study was promoted to the general audience in social media, national and local newspapers, blogs, podcasts, external channels at Norwegian University of Science and Technology (NTNU), and in national radio news.

The survey was released from eFORSK in the period August to November 2021. Participants were consecutively notified and informed through the National online health services “Helsenorge”, the digital mailbox “Digipost”, e-mail or SMS. Digitally unreachable individuals (n=42) received the survey by postal mail. One reminder was sent after two weeks to the digital responders.

### Measures

Age, gender, and habitation were automatically collected from the National Population Register through eFORSK. Sociodemographic information regarding marital status, living situation, education, profession, employment status and income were included as self-reported background information in the survey.

HRQoL was assessed by the EORTC QLQ-C30 questionnaire [7] consisting of one global health/quality of life scale, five functional scales (*physical-, role-, cognitive-, emotional- and social functioning*), 3 symptom scales (*fatigue, nausea, pain*) and 6 single items [39].

Sexual health was assessed by the EORTC sexual health questionnaire QLQ-SHQ22 [40]. It consists of eight functional scales measuring *sexual satisfaction* (among the sexually active), *importance of sexual activity* (with or without partner), *libido, impact of treatment, communication with professionals about sexual problems, insecurity with partner* (among those with partner), *femininity* (women only), *masculinity and confidence with erection* (men only), and four symptom scales assessing the impact of *sexual pain, worry about incontinence, fatigue, and vaginal dryness* (women only). The instrument has proven psychometric properties and is found applicable in research and clinical practice for assessing sexual health in both survivors and patients, across diagnosis and stages of disease, [25].

Sexual health items from the breast cancer modules QLQ-BR23 and QLQ-BR45 (not covered by the SHQ22) were added to the survey [8, 9]. These include all items in the functional scales *body image, sexual functioning, and sexual enjoyment* (in BR23/45) in addition to symptom items in the BR45-scales: *Endocrine Therapy Symptoms* and *Endocrine Sexual Symptoms* about pain or stiffness in joints or bones, pain in muscles, weight gain, mood alteration and menopausal status. As the QLQ-BR45 is not yet in Norwegian, the used items were translated by the research team according to EORTC translation manual.

Response options for all the items are *Not at all (*1) to *Very much* (4), except for the two global health / QoL items ranging from *Very poor* (1) to *Excellent* (7). To strengthen the face validity and content validity in a general population, the three items asking whether disease or treatment has impact various life conditions were given an extra response option “Not relevant”. The recall period was one week for the general health items and four weeks for the sexual items in BR23/45 and SHQ22. Scales were transformed into 0-100 scale following the EORTC scoring manual. Higher scores indicate better functioning/quality of life and higher symptom burden [41].

Morbidity was assessed by *The Self-Administered Comorbidity Questionnaire (SCQ) [39]*. The SCQ addresses the presence of up to 15 health conditions, whether the person receives treatment, and whether the condition limits any activities. In our study, morbidity was defined as having one or more morbidities that limited activities.

### Statistical analyses

Normative values are presented in six age groups (18-29, 30-39, 40-49, 50-59, 60-69 and ≥ 70 years) by means and SD, by gender and morbidity. Mean scores by gender and morbidity for all functional scales and for the most prominent symptoms were illustrated by graphs (mean, 95% CI). Group differences were tested by student t-tests. Floor and ceiling effects were calculated for the scales in QLQ-C30 and QLQ-SHQ22.

Multivariable linear regression was carried out to estimate the associations of each EORTC QLQ-C30 scale with age and gender, gender-age interaction and morbidity (0=none, 1= one or more conditions limiting activities). To predict scores for the EORTC QLQ-C30 scales for individuals or groups at a certain age and morbidity, we developed a regression model following procedures of previous publications **[5, 42]** *(Supplementary Table II)*.

### Ethics

The study was approved by the Regional Committee for Medical Research Ethics (REK 2020/58888). Study information was enclosed to the survey with completion regarded as informed consent.

## Results

### Participants

A total of 5,135 individuals responded, giving an overall response rate of 33% with almost equal gender proportion (*Table I*). The vast majority (99%) responded digitally (*Supplementary Table I)*, and participant characteristics are shown in *Table II*.

**Table I.** Response by sex and age

|  |  | **Approached**<br>**(n)** | **Answered**<br>**(n)** | **Response**<br>**rate (%)** |
| --- | --- | --- | --- | --- |
| Total |  | 15627 | 5135 | 32.86 |
| Women |  | 7900 | 2735 | 34.62 |
| Men |  | 7727 | 2400 | 31.06 |
| Age<br>group | 18-29 | 2652 | 781 | 29.45 |
|  | 30-39 | 2733 | 820 | 30.00 |
|  | 40-49 | 2462 | 876 | 35.58 |
|  | 50-59 | 2790 | 1016 | 36.42 |
|  | 60-69 | 2733 | 981 | 35.89 |
|  | 70-80 | 2257 | 661 | 29.29 |

**Table II.**
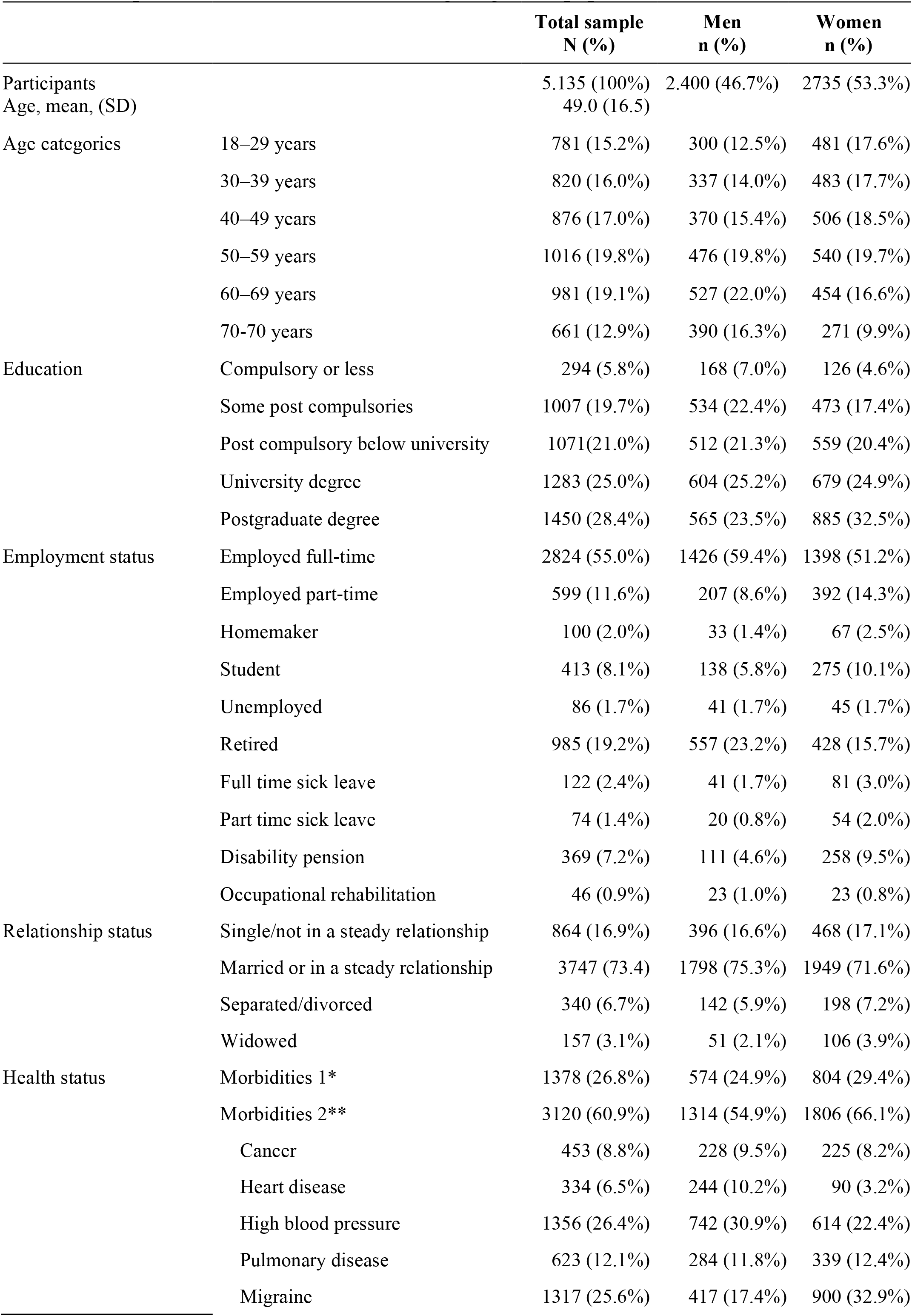
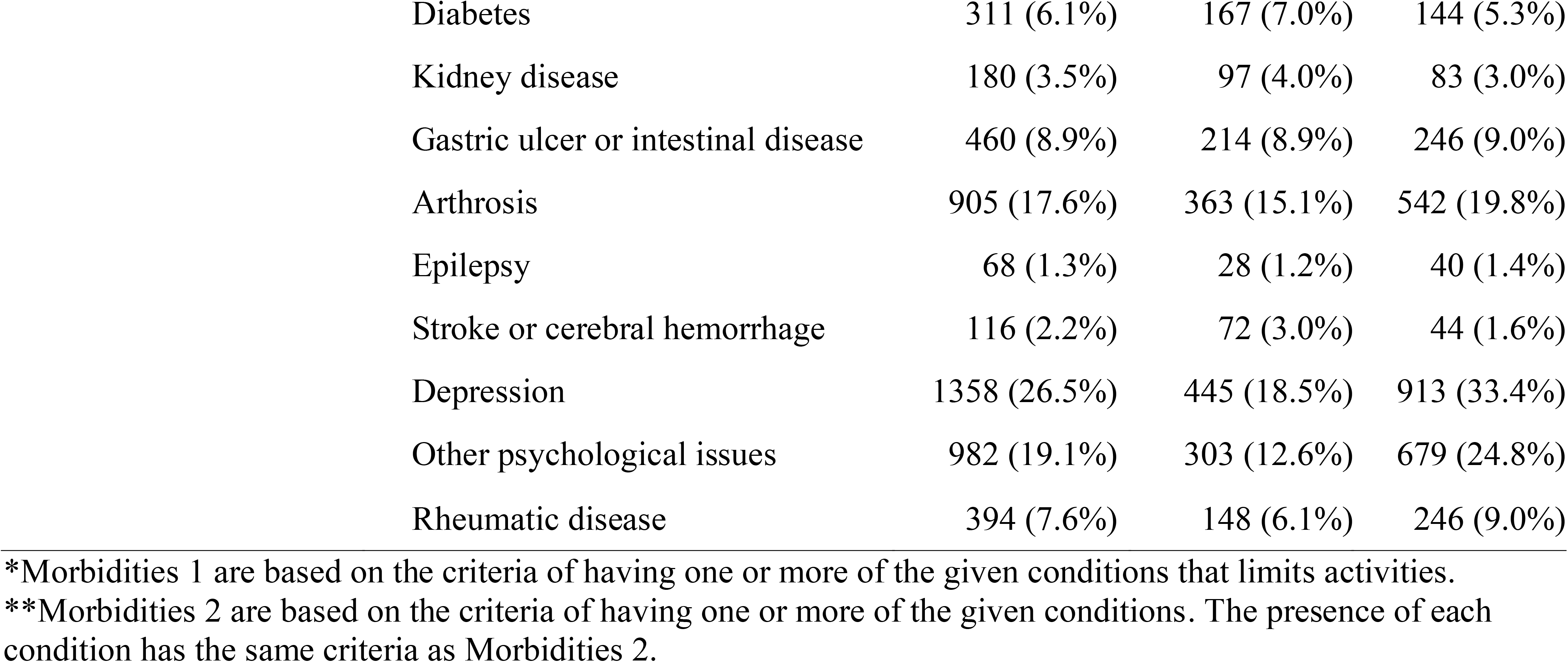
Sample characteristics for the Norwegian general population

|  |  | **Total sample**<br>**N (%)** | **Men**<br>**n (%)** | **Women**<br>**n (%)** |
| --- | --- | --- | --- | --- |
| Participants |  | 5.135 (100%) | 2.400 (46.7%) | 2735 (53.3%) |
| Age, mean, (SD) |  | 49.0 (16.5) |  |  |
| Age categories | 18–29 years | 781 (15.2%) | 300 (12.5%) | 481 (17.6%) |
|  | 30–39 years | 820 (16.0%) | 337 (14.0%) | 483 (17.7%) |
|  | 40–49 years | 876 (17.0%) | 370 (15.4%) | 506 (18.5%) |
|  | 50–59 years | 1016 (19.8%) | 476 (19.8%) | 540 (19.7%) |
|  | 60–69 years | 981 (19.1%) | 527 (22.0%) | 454 (16.6%) |
|  | 70–70 years | 661 (12.9%) | 390 (16.3%) | 271 (9.9%) |
| Education | Compulsory or less | 294 (5.8%) | 168 (7.0%) | 126 (4.6%) |
|  | Some post compulsories | 1007 (19.7%) | 534 (22.4%) | 473 (17.4%) |
|  | Post compulsory below university | 1071(21.0%) | 512 (21.3%) | 559 (20.4%) |
|  | University degree | 1283 (25.0%) | 604 (25.2%) | 679 (24.9%) |
|  | Postgraduate degree | 1450 (28.4%) | 565 (23.5%) | 885 (32.5%) |
| Employment status | Employed full-time | 2824 (55.0%) | 1426 (59.4%) | 1398 (51.2%) |
|  | Employed part-time | 599 (11.6%) | 207 (8.6%) | 392 (14.3%) |
|  | Homemaker | 100 (2.0%) | 33 (1.4%) | 67 (2.5%) |
|  | Student | 413 (8.1%) | 138 (5.8%) | 275 (10.1%) |
|  | Unemployed | 86 (1.7%) | 41 (1.7%) | 45 (1.7%) |
|  | Retired | 985 (19.2%) | 557 (23.2%) | 428 (15.7%) |
|  | Full time sick leave | 122 (2.4%) | 41 (1.7%) | 81 (3.0%) |
|  | Part time sick leave | 74 (1.4%) | 20 (0.8%) | 54 (2.0%) |
|  | Disability pension | 369 (7.2%) | 111 (4.6%) | 258 (9.5%) |
|  | Occupational rehabilitation | 46 (0.9%) | 23 (1.0%) | 23 (0.8%) |
| Relationship status | Single/not in a steady relationship | 864 (16.9%) | 396 (16.6%) | 468 (17.1%) |
|  | Married or in a steady relationship | 3747 (73.4) | 1798 (75.3%) | 1949 (71.6%) |
|  | Separated/divorced | 340 (6.7%) | 142 (5.9%) | 198 (7.2%) |
|  | Widowed | 157 (3.1%) | 51 (2.1%) | 106 (3.9%) |
| Health status | Morbidities 1* | 1378 (26.8%) | 574 (24.9%) | 804 (29.4%) |
|  | Morbidities 2** | 3120 (60.9%) | 1314 (54.9%) | 1806 (66.1%) |
|  | Cancer | 453 (8.8%) | 228 (9.5%) | 225 (8.2%) |
|  | Heart disease | 334 (6.5%) | 244 (10.2%) | 90 (3.2%) |
|  | High blood pressure | 1356 (26.4%) | 742 (30.9%) | 614 (22.4%) |
|  | Pulmonary disease | 623 (12.1%) | 284 (11.8%) | 339 (12.4%) |
|  | Migraine | 1317 (25.6%) | 417 (17.4%) | 900 (32.9%) |

|  |  |  |  |
| --- | --- | --- | --- |
| Diabetes | 311 (6.1%) | 167 (7.0%) | 144 (5.3%) |
| Kidney disease | 180 (3.5%) | 97 (4.0%) | 83 (3.0%) |
| Gastric ulcer or intestinal disease | 460 (8.9%) | 214 (8.9%) | 246 (9.0%) |
| Arthrosis | 905 (17.6%) | 363 (15.1%) | 542 (19.8%) |
| Epilepsy | 68 (1.3%) | 28 (1.2%) | 40 (1.4%) |
| Stroke or cerebral hemorrhage | 116 (2.2%) | 72 (3.0%) | 44 (1.6%) |
| Depression | 1358 (26.5%) | 445 (18.5%) | 913 (33.4%) |
| Other psychological issues | 982 (19.1%) | 303 (12.6%) | 679 (24.8%) |
| Rheumatic disease | 394 (7.6%) | 148 (6.1%) | 246 (9.0%) |

### Normative data for EORTC QLQ-C30

Normative scores for the EORTC QLQ-C30 scales are presented in *Table III*. The most prominent symptoms were fatigue, insomnia, and pain. Floor and ceiling effects are displayed in *Supplementary Table II*. The regression model for predicting individual EORTC QLQ-C30 normative scores for age-, sex- and morbidity is provided in *Table IV*.

**Table III.**
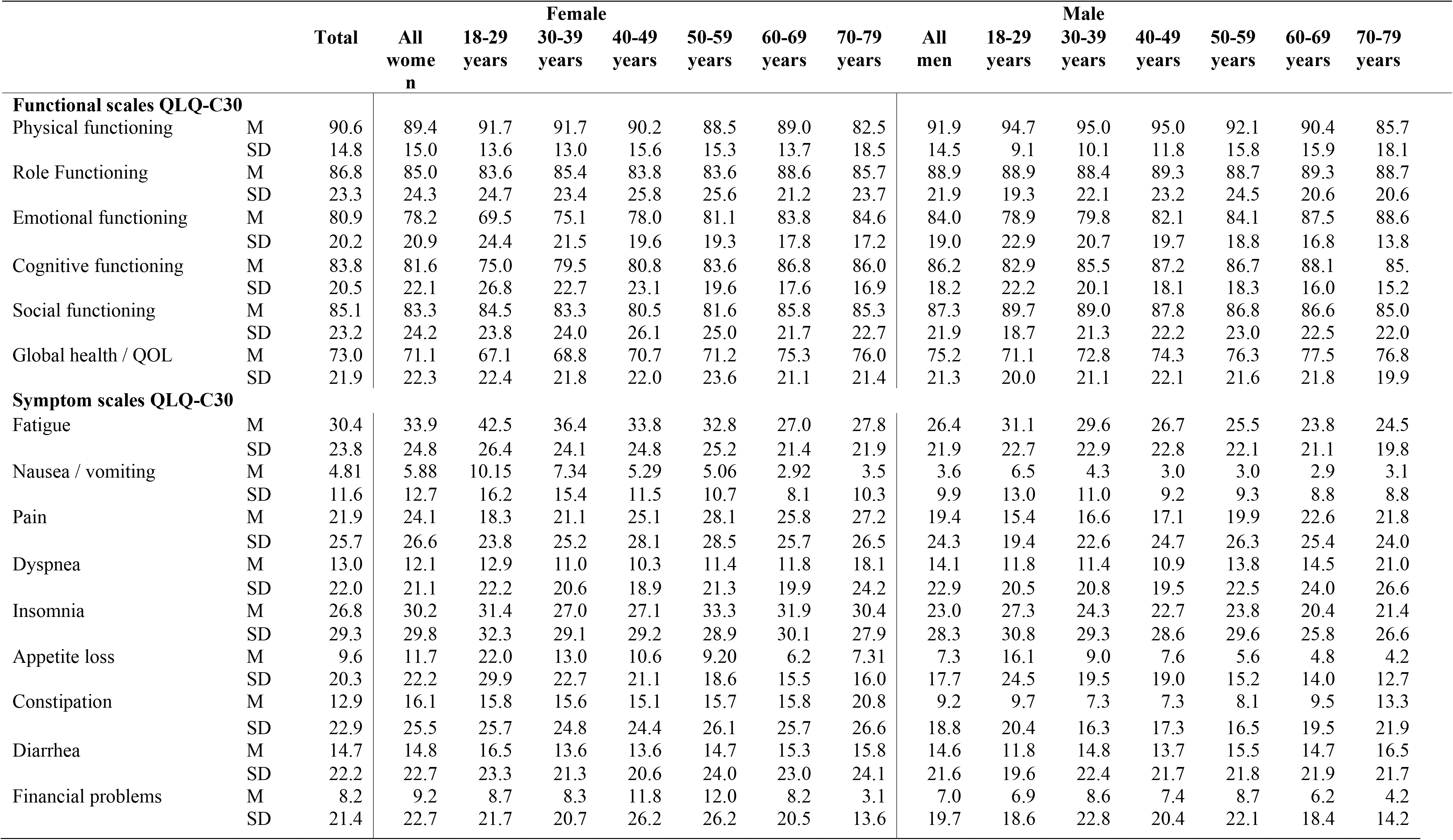
Reference values for EORTC QLQ-C30 by sex and age groups

**Table IV:**
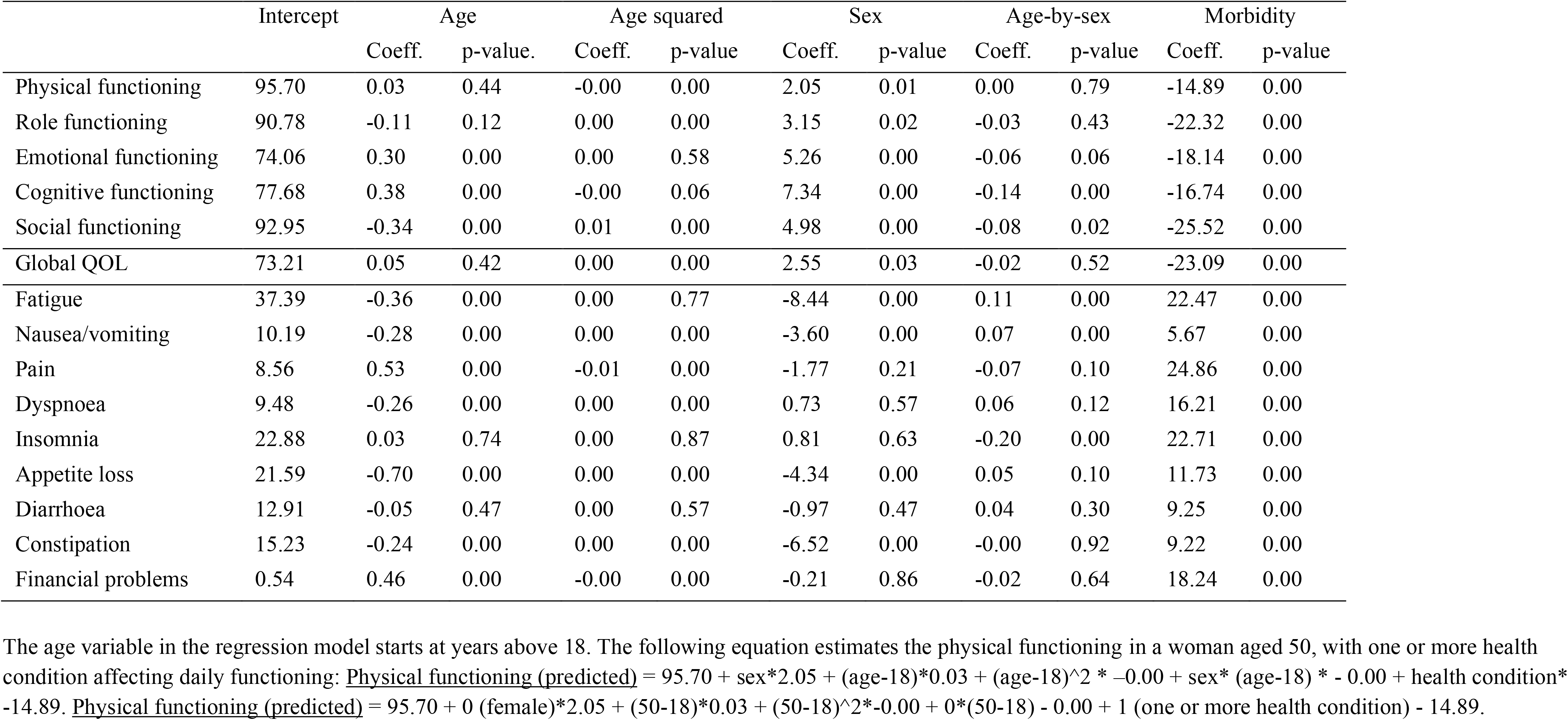
Regression model for the EORTC QLQ-C30 values by age, sex and morbidity

Women reported generally lower functioning and higher symptom scores than men. Gender differences were most pronounced for emotional functioning, pain, fatigue, and insomnia. The youngest women (18-29 years) reported poorer emotional function (9.4 points) and more fatigue (11.4 point), and the older women (59-79 years) reported more pain (8.2 points) and sleep problems (9.5 point) compared to men in the corresponding age groups *(Figure I, Table III)*.

**Figure IA.**
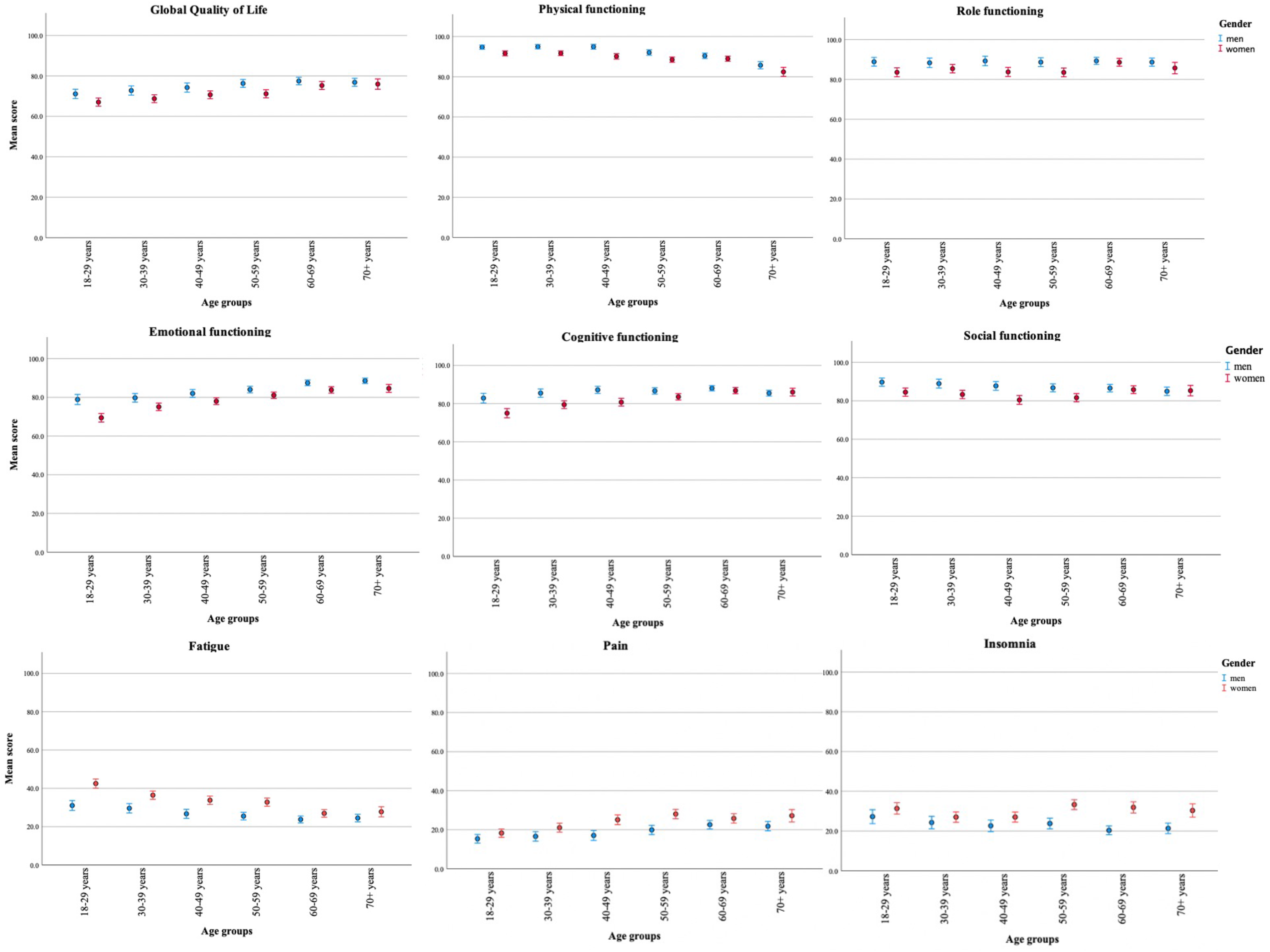
EORTC QLQ-C30 - Mean scores of all the functional scales and the most prominent symptoms for men and women in the Norwegian general population, presented in 10-years age groups from 29-79 years. Error bars represent mean scores with 95% confidence intervals. Higher scores on functional scales indicate better functioning and higher scores on symptom scales implies more symptom burden.

**Figure IB.**
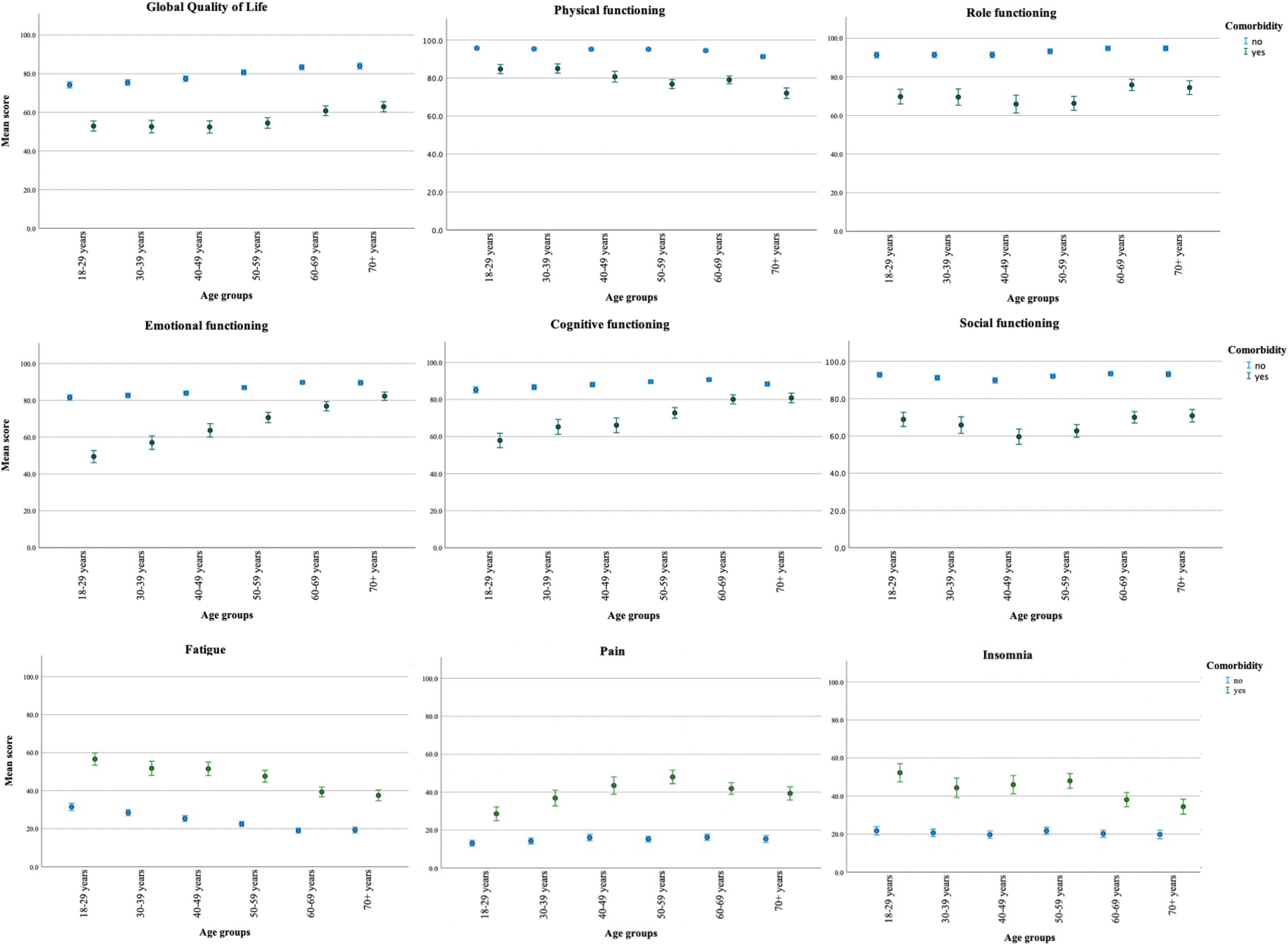
EORTC QLQ-C30 - Mean scores of all the functional scales and the most prominent symptoms for persons with and without morbidities in the Norwegian general population, presented in 10-years age groups from 29-79 years. Morbidity is based on the criteria of having one or more conditions that causes limitations in activity. Error bars represent mean scores with 95% confidence intervals. Higher scores on functional scales indicate better functioning and higher scores on symptom scales implies more symptom burden.

Respondents with morbidities scored significantly lower on global QOL and functional scales and higher on symptom scales than persons without morbidities across all age-groups, *Figure II*. The largest differences were observed among the youngest (18-29 years) on emotional and cognitive functioning (32 and 28 points). Physical functioning was most divergent in the middle aged (15 points) and oldest age group (19 points).

**Figure IIA.**
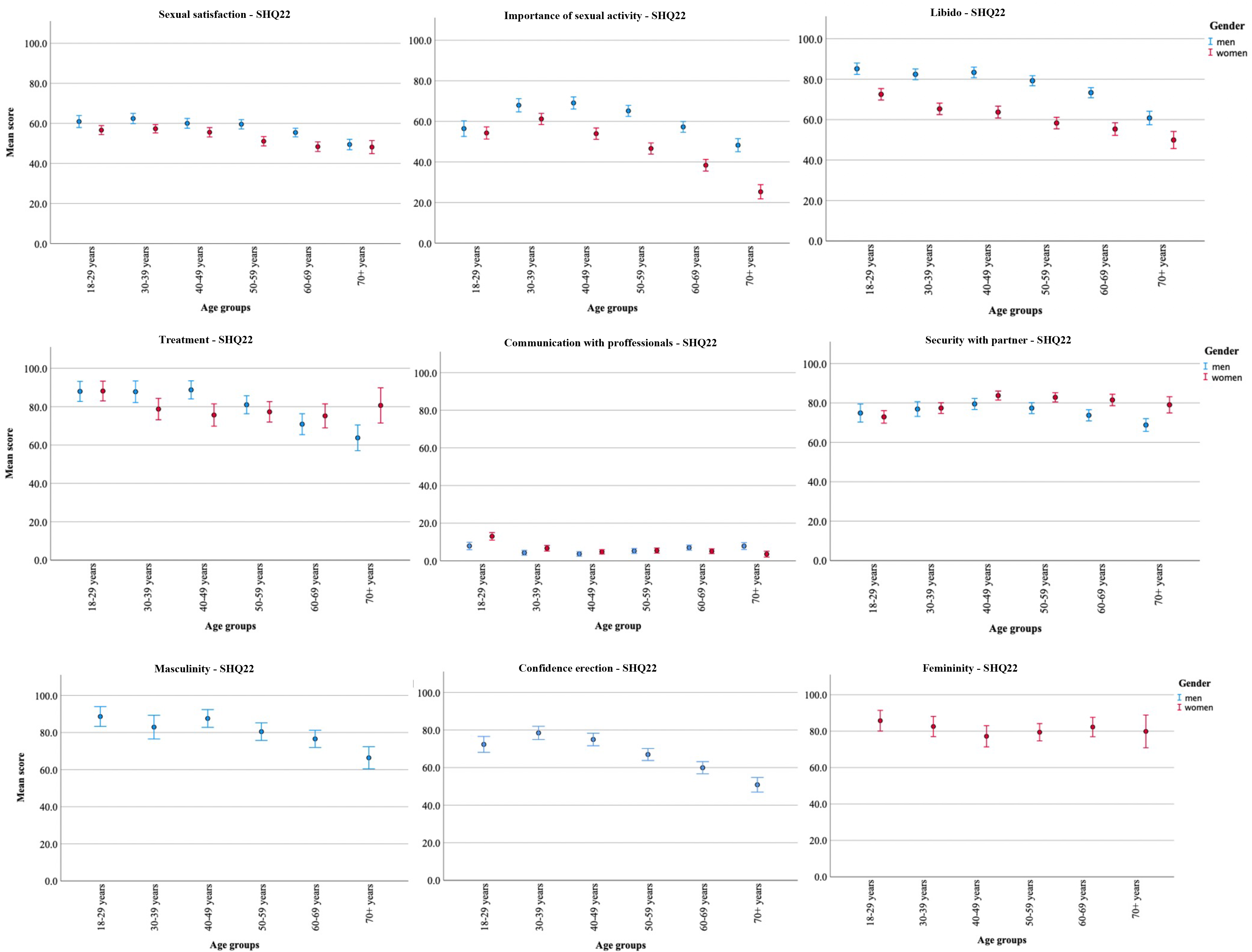
EORTC QLQ-SHQ22, QLQ-BR23 and QLQ-BR45 - Mean scores of functional and symptom scales for men and women in the Norwegian general population, presented in 10-years age groups from 29-79 years. Error bars represent mean scores with 95% confidence intervals. Higher scores on functional scales indicate better functioning and higher scores on symptom scales implies more symptom burden. For QLQ-SHQ22 all scales are presented, for QLQ-BR23/45 only scales applicable for the general population are included.

**Figure IIB.**
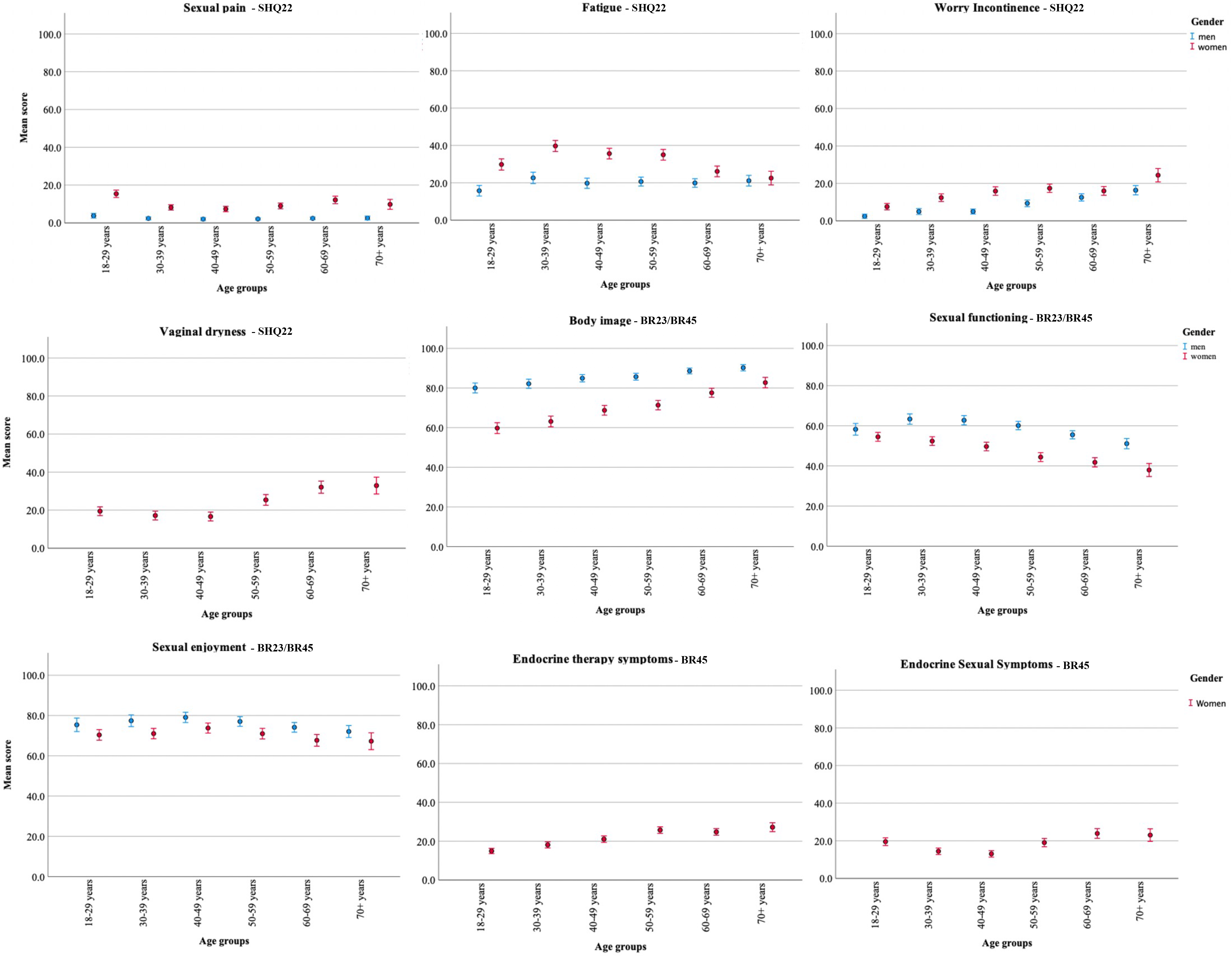
EORTC QLQ-SHQ22, QLQ-BR23 and QLQ-BR45 - Mean scores of functional and symptom scales for persons with and without morbidities in the Norwegian general population, presented in 10-years age groups from 29-79 years. Morbidity is based on the criteria of having one or more conditions that causes limitations in activity. Error bars represent mean scores with 95% confidence intervals. Higher scores on functional scales indicate better functioning and higher scores on symptom scales implies more symptom burden. For QLQ-SHQ22 all scales are presented, for QLQ-BR23/45 only scales applicable for the general population are included.

Among symptoms, *Insomnia and Fatigue* displayed the largest differences between persons with and without morbidities among the youngest (25 and 31 points) and middle-aged groups (26 points on both symptoms), whereas pain differences were largest in the middle-aged group (28 points). All group differences were highly significant (p<0.001).

### Normative data for EORTC QLQ-SHQ22 and the sexual scales in EORTC QLQ-BR23 and QLQ-BR45

Normative scores for the EORTC QLQ-SHQ22 and sexual scale scores from the EORTC QLQ-BR23/BR45 are presented in *Table V*. Floor and ceiling effects are displayed in *Supplementary Table III*. Among symptoms *Fatigue* influenced sexual life most (*Figure III, Table V*).

**Table IV.**
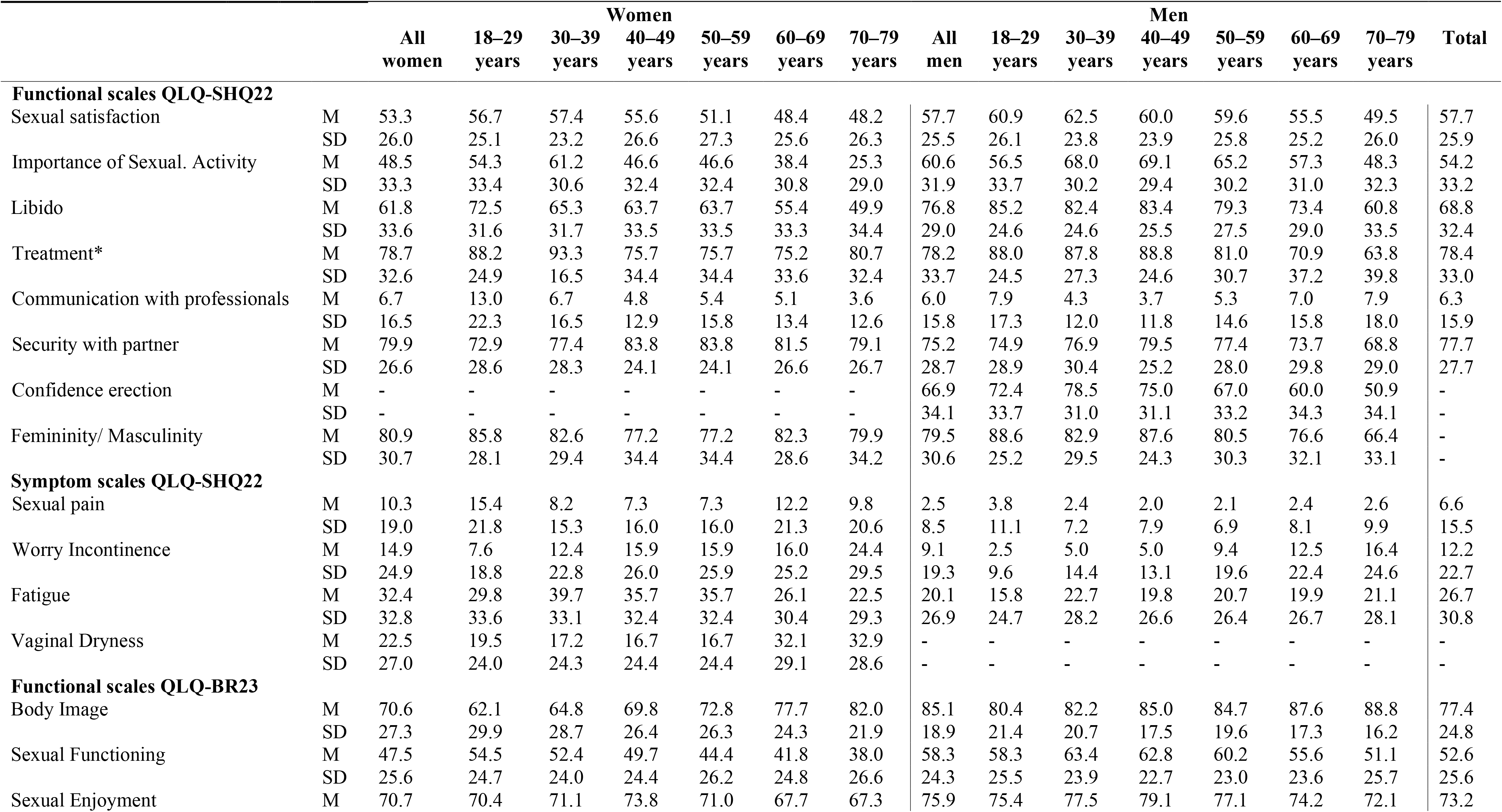
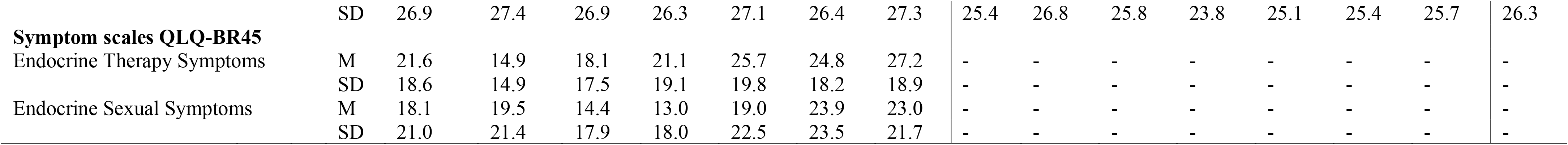
Reference values for EORTC QLQ-SHQ22 and for the sexual scales in QLQ-BR23 and QLQ-BR45

Women reported generally lower functioning and higher symptom scores than men (*Figure IV)*. Gender differences in the functional scales were most pronounced for *Importance of Sexual Activity* (22 points), *Libido* (20 points), *Body Image* particularly among the youngest (20 points, 18-29 years), and *Sexual Functioning* which differed by 10-13 points between genders from the age of 30-69 years.

The influence of *Fatigue* on sexual life was most gender divergent among the youngest (18-39 years) and middle-aged groups (40-59 years) with a gap between 14 and 17 points *(Figure III and Table V)*.

Respondents with morbidities scored in general lower on functioning and higher on symptoms than persons without morbidities, as illustrated in *Figure IV*. The largest mean difference was observed in men with and without comorbidities on masculinity aged 40-49 years with a gap of 30 points. In both genders *Sexual Satisfaction, Libido, Security with partner, Body Image* and *Sexual Enjoyment* were significantly lower in persons with morbidities with most pronounced differences among the youngest and middle-aged. *Treatment* had significantly more impact on sexual life in persons with morbidities, and the youngest men (18-39 years) with morbidities had significantly less *Confident in erection*.

The influence of *Fatigue* on sexual life displayed the largest differences among the youngest and the middle-aged group (26 points, 18-49 years) between persons with and without morbidities (*Figure IV*). Among women, *Endocrine Therapy Symptoms* (which reflected mood swings, pain or stiffness joints, bones and muscles and weight gain) was most divergent in the middle-aged group (19 points) and *Femininity* in the age group 60-60 years (21 points). Group differences were all highly significant (p<0.001).

## Discussion

The present study provides new Norwegian general population normative values on the EORTC QLQ-C30 and is the first study to present normative values on the EORTC sexual health questionnaire QLQ-SHQ22, and the sexual scales in QLQ-BR23 and QLQ-BR45.

Global health/QoL, emotional and cognitive functioning increased by age in the Norwegian population, in line with recent studies in Italy [5] and Australia [43], but in contrast to previous normative studies in Europe [32, 34, 36, 44]. This pattern could be due to an improved health care system and a healthier lifestyle among present elderly citizens.

The most prominent symptoms were fatigue, insomnia, and pain, similar to the Italian [5], Swedish [28], Danish [19], and the previous Norwegian normative population samples [30, 32]. However, compared to prior EORTC studies, a new symptom distribution across age-groups was observed where fatigue and insomnia were more severe among the youngest age groups, and particularly among women. Increased fatigue among the youngest was also found in recent Norwegian normative studies of generic HRQoL questionnaires [45, 46], and similar trends of higher symptom burden among the youngest has been found in the latest European normative studies [5, 20, 27]. This pattern may be explained by high influence of social media, high demands and often many options to navigate between among youths of today.

Norwegian women reported more symptoms and lower functioning than men, in line with previous studies [5, 17, 19, 20, 27-29]. Gender differences were more prominent in the Norwegian population than in the German [47] and Danish populations[19], but in line with findings in recent Italian study [5]. However, the distinction in score patterns between groups defined by having morbidity or not, were far more pronounced than sex differences, and congruent to other norm studies with valid detection of morbidities [5, 19, 20, 27, 48].

Normative scores for sexual health are the first of its kind; for EORTC QLQ-SHQ22 worldwide and for BR-23/45 in Scandinavia, though such knowledge is highly required both in research and clinical use [24]. Gender differences were even more pronounced in the sexual dimensions of HRQOL, particularly among the youngest age groups where females scored lower. The most striking result was the huge gender gap in the functional scales *Body Image, Libido, Sexual Functioning*, and *Importance of Sexual Activity*. Among the symptom scales, the impact of *Fatigue* on sex life was most prominent in the age groups from 30-59 and may explain some of the low sexual functioning scores in these age groups. Morbidities had a generally negative impact on all sexual domains, except for the importance of sexual activity. The Norwegian general population report nearly no communication with healthcare professionals about sexual topics, in line with frequently described barriers in the patient-clinician interaction around sexual health [49, 50].

Limitations of the present study could be the response rate of 35%, which might threaten the generalizability to the general population. However, adding questions about sensitive topics implies an increased risk of lower response rate [24]. We tried to counteract this through several efforts in the data collection process. The response rate, 35% for women and 31% for men, was evidently lower than in panel data studies [20, 24, 31], however beyond expected in a Norwegian general population sample [51], and similar to the last updated reference data on EORTC in Norway [30].The new breast module QLQ-BR45 was not available in Norwegian, and our translation of specific items could be suboptimal.

Strengths of this study are the large sample size and the high representativity to the general population. We performed age- and gender stratified random sampling from the Norwegian population and included a large sample ensuring high statistical power in subgroup analyses. Following advice from previous studies [5, 27], morbidity was registered in more detail by adding information on its influence on daily functioning [39]. Our final national sample of 5,135 participants is the largest normative EORTC study in Europe and is the first to include sexual health as an important aspect of HROoL.

## Conclusion

The present study presents updated Norwegian normative general population data for the EORTC QLQ-C30 and is the first to provide normative values for the EORTC sexual health questionnaire QLQ-SHQ22 and the sexual subscales of the breast modules QLQ-BR23 and QLQ-BR45, for all separately in age groups by gender and morbidity. Normative values can serve as a support when interpreting HRQoL profiles in Norwegian cancer populations.

## Data Availability

All data produced in the present study are available upon reasonable request to the authors

## Availability of data and materials

The data are available on request to the corresponding author.

---

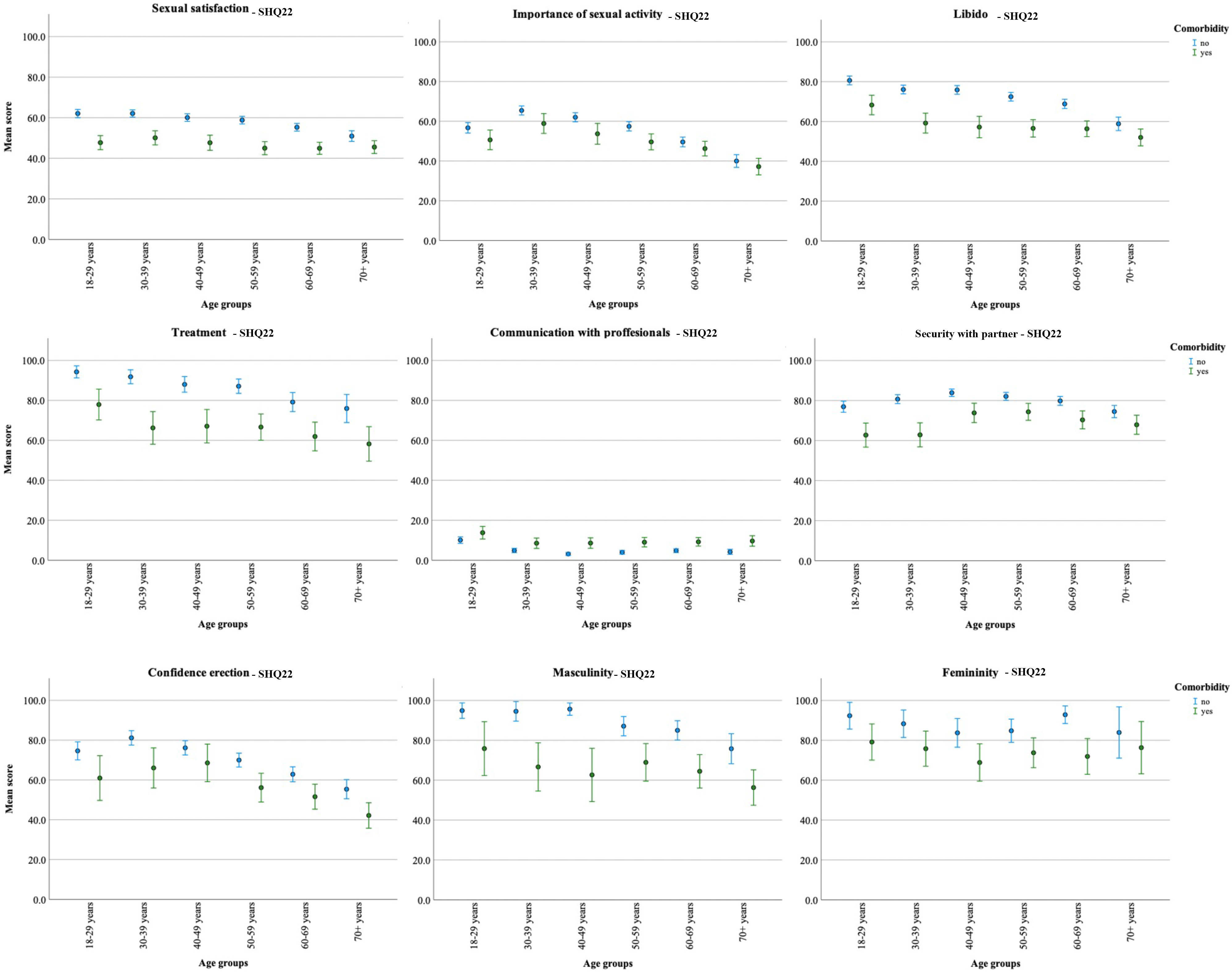

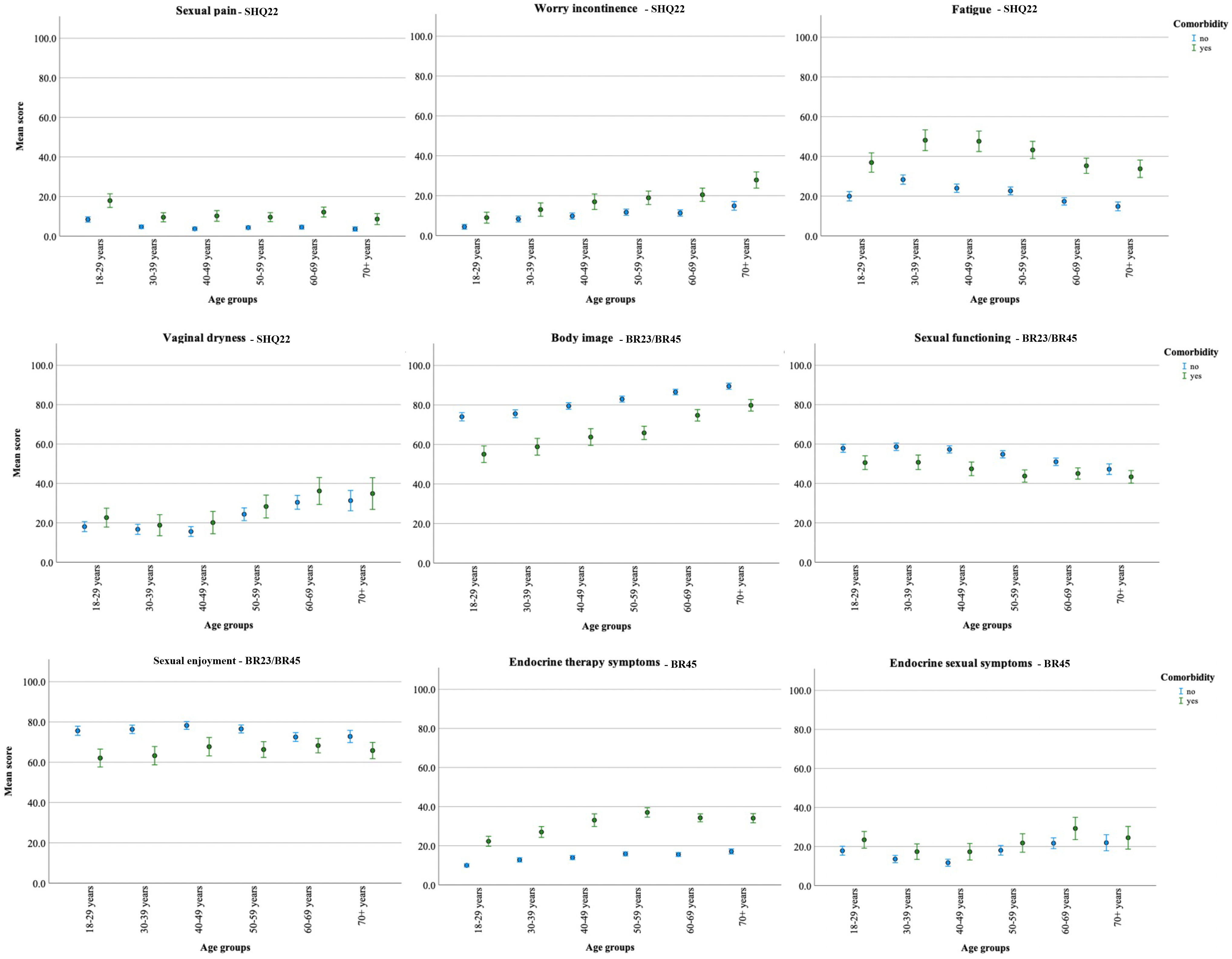

**Supplementary Table I:** Platform for participation in the survey

| Participation<br>by | **Total sample**<br>N=5135 | **Men**<br>n=2400 | **Women**<br>n=2735 |
| --- | --- | --- | --- |
| National online health services | 4961 (96.6%) | 2305 (96.0%) | 2656 (97.1%) |
| DigiPost (digital postbox) | 61 (1.2%) | 35 (1.5%) | 26 (1.0%) |
| E-mail / Text message | 71 (1.4%) | 40 (1.7%) | 31 (1.1%) |
| Paper schemes by postal mail | 42 (0.8%) | 20 (0.8%) | 22 (0.8%) |

**Supplementary Table II.** Floor and ceiling effect in the EORTC QLQ-C30 scales

|  | **Lowest**<br>**possible**<br>**scores**<br>**0 points**<br>**(%)** | **Highest**<br>**possible**<br>**scores**<br>**100 points**<br>**(%)** |
| --- | --- | --- |
| Physical functioning | 0.0 | 50.0 |
| Role functioning | 1.7 | 67.3 |
| Emotional functioning | 0.4 | 30.8 |
| Cognitive functioning | 0.9 | 45.8 |
| Social functioning | 1.4 | 61.2 |
| Global QLQ | 0.5 | 18.2 |
| Fatigue | 17.8 | 1.8 |
| Nausea/Vomiting | 80.5 | 0.1 |
| Pain | 42.4 | 2.6 |
| Dyspnea | 69.2 | 1.5 |
| Insomnia | 44.6 | 5.8 |
| Appetite loss | 78.2 | 1.2 |
| Constipation | 71.0 | 2.0 |
| Diarrhea | 64.2 | 1.4 |
| Financial problems | 84.3 | 2.5 |

**Supplementary Table III.** Floor and ceiling effect in the EORTC QLQ-SHQ22

|  | **Lowest**<br>**possible**<br>**scores**<br>**0 points**<br>**(%)** | **Highest**<br>**possible**<br>**scores**<br>**100 points**<br>**(%)** |
| --- | --- | --- |
| Sexual satisfaction | 2.5 | 2.6 |
| Importance of sexual activity | 16.0 | 21.8 |
| Libido | 9.7 | 41.0 |
| Treatment | 8.5 | 64.2 |
| Communication with professionals | 84.1 | 0.5 |
| Security with partner | 4.4 | 52.2 |
| Confidence erection | 11.3 | 41.3 |
| Femininity/Masculinity | 6.4 | 66.5 |
| Masculinity | 6.0 | 62.6 |
| Sexual pain | 76.9 | 0.4 |
| Worry Incontinence | 73.0 | 2.0 |
| Fatigue | 47.7 | 7.1 |
| Vaginal dryness | 51.0 | 3.3 |

## Notes

### Competing Interest Statement

The authors have declared no competing interest.

### Funding Statement

This study was funded by the Norwegian Cancer Society, the Norwegian Breast Cancer Society and The Norwegian University of Science and Technology

### Author Declarations

The study was approved by the Ethics Committee of Central Norway, Regional Committee for Medical Research Ethics (REK numbers 2020/58888). Study information was enclosed to the survey with completion regarded as informed consent.

